# Association of Insurance Payor with Time to Discharge to Inpatient Rehabilitation After Ischemic Stroke

**DOI:** 10.64898/2026.07.08.26357596

**Authors:** Rajit J. Shah, Benjamin King, Sarah Strobel, Rolake Feyisetan

## Abstract

**Background:** Transition timing to post-acute rehabilitation after ischemic stroke is heavily influenced by non-clinical factors, introducing potential systemic disparities in care access. We evaluated the association between insurance payor status and acute hospital length of stay (LOS) prior to inpatient rehabilitation discharge among critically ill stroke patients.

**Methods:** Using the MIMIC-IV database, we identified ICU-admitted adults with ischemic stroke discharged to inpatient rehabilitation (n=1,285). The primary outcome was hospital LOS prior to rehab transfer. Multivariable log-transformed linear regression evaluated the association with insurance payor (Medicare, private, other/unknown; reference: Medicaid), adjusting for demographics, diagnostic-code counts (medical complexity), and ICU LOS (acute illness severity).

**Results:** Median hospital LOS before rehab discharge was longest for Medicaid patients (13.2 days) compared with private insurance (11.0 days) and Medicare (9.5 days). In the adjusted model, Medicare insurance was associated with a significantly shorter transition time to inpatient rehabilitation, corresponding to a 13.5% shorter acute hospital stay (adjusted LOS ratio 0.87; 95% CI: 0.79–0.96; p=0.005) relative to Medicaid. Private insurance demonstrated a descriptive trend toward shorter LOS that did not achieve statistical significance (adjusted LOS ratio 0.93; 95% CI: 0.84–1.02; p=0.122). Other and unknown payor categories showed no significant differences.

**Conclusions:** Insurance payor status serves as an independent predictor of acute care transition timing for stroke patients requiring inpatient rehabilitation. The prolonged acute stays observed among Medicaid beneficiaries suggest significant non-clinical, administrative bottlenecks in post-acute placement, underscoring the critical need for standardized, streamlined insurance approval pathways to ensure equitable neurological recovery.

**Perspective:** What New Question Does This Study Raise?

- This study highlights that even when adjusting for intensive care utilization and underlying medical complexity, public insurance status independently correlates with prolonged acute hospital stays prior to inpatient rehabilitation transfer, raising critical questions regarding the exact administrative mechanisms and pre-authorization workflows driving these non-clinical bottlenecks.

What Question Should Be Addressed Next?

- Future investigations must evaluate whether these insurance-driven operational delays in acute care transitions directly translate to inferior long-term functional recovery, higher stroke recurrence, or increased 30-day readmission rates for vulnerable patient cohorts.
- Additionally, research should investigate how these acute-phase disposition delays cascade upstream to affect emergency department boarding metrics, ambulance gridlock, and the overall operational costs absorbed by safety-net hospital systems.

## Introduction

Stroke remains a leading public health challenge in the United States, currently ranking as the fourth leading cause of death and a driver of long-term adult disability. Current data show an annual incidence of 800,000 new or recurrent strokes, with projections suggesting that one in four adults over age 25 will experience a stroke in their lifetime.^1^ This burden is exacerbated by significant gaps in healthcare coverage. Among working-age stroke survivors, nearly 14% are uninsured, while over 35% are classified as underinsured.^1^ These coverage gaps directly correlate with a lack of management for co-morbid risk factors such as hypertension and diabetes, leading to a 7% increase in age-adjusted stroke mortality rates over the last decade.^1^

The impact of insurance status extends beyond prevention, influencing acute intervention and resource utilization. Uninsured patients are 10% less likely to receive timely thrombolytic therapy compared to privately insured patients, a disparity that contributes to a 56% higher in-hospital mortality rate for acute ischemic stroke.^2^ Furthermore, the economic burden of recovery is immense, with first-year medical costs averaging $70,601 per patient, the majority of which is driven by rehabilitation.^1^ For patients with complex needs, these costs can scale significantly.

Acute ischemic stroke management necessitates a transition from acute stabilization to post-acute care (PAC). While clinical severity is a primary driver of recovery, the economic and systemic burdens associated with stroke can dictate patient recovery trajectory.^3^ Discharge timing and destination are multidimensional processes influenced heavily by non-clinical factors, most notably insurance payor status. Evidence from large-scale registries, such as the Get With The Guidelines-Stroke program, has demonstrated that insurance status is a significant predictor of outcomes and the use of rehabilitation services.^4,5^ These disparities are particularly pronounced among working-age stroke survivors, where a lack of private insurance often correlates with reduced access to high-intensity institutional PAC.^6–8^

The type of government-sponsored coverage also plays a critical role. For older adults (aged 65+), dual eligibility for both Medicare and Medicaid serves as a significant marker for increased medical complexity and socioeconomic disadvantage, often resulting in higher initial stroke severity. Furthermore, these dually enrolled patients face substantial disparities in post-acute care, as they are significantly less likely to be discharged to high-quality skilled nursing facilities or home health agencies compared to those with traditional Medicare alone.^9^ Recent data also indicates substantial disparities in post-acute care quality between Medicare Advantage and traditional Medicare beneficiaries.^9^ These inequities are not limited to stroke as similar associations between payor source and rehabilitation outcomes have been observed in traumatic brain injury populations.^10^ Moreover, these financial barriers often intersect with racial, ethnic, and regional disparities, creating a compounding effect on service utilization.^11^

Despite the growing body of literature on general rehabilitation access, there remains a need to investigate how insurance payor specifically impacts the temporal aspects of acute recovery, such as hospital length of stay and time to discharge to inpatient rehabilitation among ICU ischemic stroke patients. Data from the MIMIC-IV database suggests that Medicaid and “No Charge” status are independent predictors of prolonged lengths of stay, highlighting how payor status acts as a systemic bottleneck that delays transition to sub-acute care facilities.^12^ Utilizing the MIMIC-IV database, a comprehensive and freely accessible electronic health record dataset, this study aims to elucidate the relationship between insurance payor and discharge timing in a critically ill ischemic stroke cohort. Understanding these dynamics is essential for identifying systemic bottlenecks and informing policy that ensures equitable transitions of care for all patients, regardless of their healthcare coverage.

## Methods

We conducted a retrospective cohort study using the MIMIC-IV v3.1 database to evaluate the association between insurance payor and time to discharge to inpatient rehabilitation among ICU-admitted adults with ischemic stroke. MIMIC-IV is a freely accessible, de-identified electronic health record database containing real-world clinical data from hospital and ICU admissions at Beth Israel Deaconess Medical Center. Adult ICU admissions with qualifying ischemic stroke ICD-9/ICD-10 diagnosis codes were identified and linked with demographic, administrative, ICU stay, medication, and discharge data.

The source cohort included 4,300 ICU-admitted ischemic stroke patients, of whom 1,285 (29.9%) were discharged to inpatient rehabilitation and comprised the primary analytic cohort. The primary outcome was hospital length of stay prior to inpatient rehabilitation discharge, calculated as the difference between hospital admission time and discharge time among patients discharged to inpatient rehabilitation. Insurance payor was the primary exposure and was categorized as Medicare, Medicaid, private insurance, other, or unknown. Race/ethnicity was collapsed into White, Black, Asian, Hispanic, and Other.

Among the inpatient rehabilitation discharge cohort, the most common insurance categories were Medicare (n=791), private insurance (n=317), and Medicaid (n=150). Median length of stay before inpatient rehabilitation discharge was 9.5 days for Medicare, 11.0 days for private insurance, and 13.2 days for Medicaid.

The primary subgroup analysis used multivariable log-transformed length-of-stay regression to estimate the association between insurance payor and time to inpatient rehabilitation discharge. Models adjusted for age, sex, collapsed race/ethnicity, diagnosis-code count as a proxy for medical complexity, ICU length of stay as a proxy for acute illness severity, tPA receipt, and tPA timing when available. Medicaid was used as the reference insurance category. Model results were expressed as adjusted length-of-stay ratios and percent differences in LOS relative to Medicaid. Robust standard errors were used to account for heteroscedasticity. The broader ICU ischemic stroke cohort was summarized descriptively to characterize overall discharge disposition patterns.

## Results

A total of 4,300 ICU-admitted patients meeting the diagnostic criteria for acute ischemic stroke were identified within the primary database, of whom 1,285 (29.9%) were explicitly discharged to inpatient rehabilitation and comprised the primary analytic cohort. The adjusted coefficients and multi-variable ratios for this model are detailed in Table 1. The baseline distribution of insurance payor, race/ethnicity, and inpatient rehabilitation discharge status across the broader study population is detailed comprehensively in Table 2. The complete breakdown of discharge disposition pathways across each distinct insurance payor group highlights key variations in care transitions (Figure 4). Within this post-acute transition cohort, the distribution of primary insurance payors was heavily weighted toward Medicare (n=791), followed by private insurance (n=317) and Medicaid (n=150). Unadjusted descriptive analysis revealed that the median hospital length of stay (LOS) prior to inpatient rehabilitation transfer was longest for Medicaid beneficiaries (13.2 days) compared with privately insured patients (11.0 days) and Medicare beneficiaries (9.5 days).

**Table 1.**
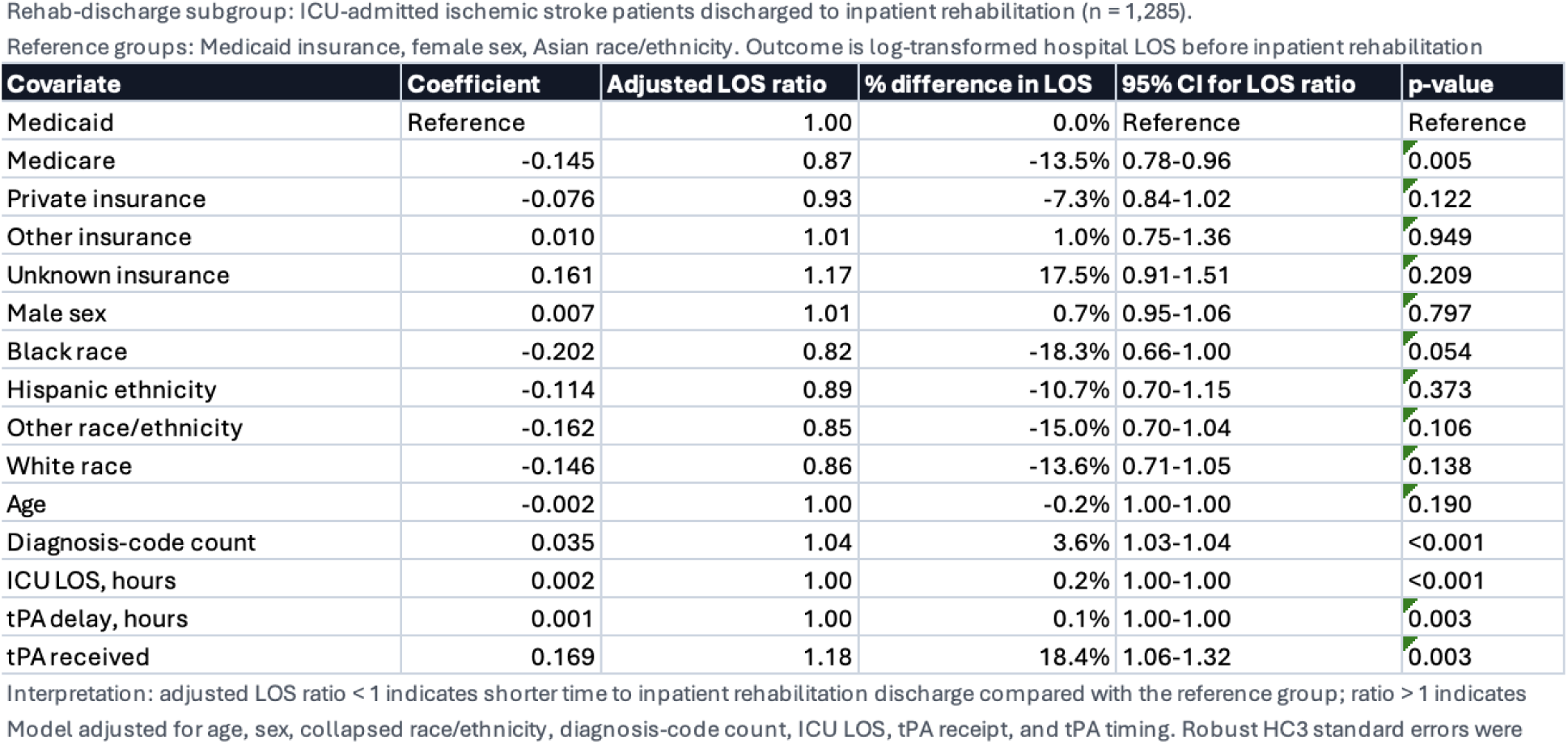
Adjusted association between insurance payor and time to inpatient rehabilitation discharge after ischemic stroke. This multivariable log-transformed length-of-stay regression model evaluated the association between insurance payor and time from hospital admission to discharge to inpatient rehabilitation among ICU-admitted ischemic stroke patients who were discharged to inpatient rehabilitation. The model adjusted for age, sex, collapsed race/ethnicity, diagnosis-code count as a proxy for medical complexity, ICU length of stay as a proxy for acute illness severity, tPA receipt, and tPA timing when available. Medicaid was used as the reference insurance category. Compared with Medicaid, Medicare insurance was significantly associated with shorter time to inpatient rehabilitation discharge, corresponding to a 13.5% shorter adjusted LOS (adjusted LOS ratio 0.87; p=0.005). Private insurance showed a similar but non-significant trend toward shorter LOS (adjusted LOS ratio 0.93; p=0.122). The overall model was statistically significant (p<0.001) and explained approximately 56.9% of the variance in log-transformed LOS (R²=0.569).

| Covariate | Coefficient | Adjusted LOS ratio | % difference in LOS | 95% CI for LOS ratio | p-value |
| --- | --- | --- | --- | --- | --- |
| Medicaid | Reference | 1.00 | 0.0% | Reference | Reference |
| Medicare | -0.145 | 0.87 | -13.5% | 0.78-0.96 | 0.005 |
| Private insurance | -0.076 | 0.93 | -7.3% | 0.84-1.02 | 0.122 |
| Other insurance | 0.010 | 1.01 | 1.0% | 0.75-1.36 | 0.949 |
| Unknown insurance | 0.161 | 1.17 | 17.5% | 0.91-1.51 | 0.209 |
| Male sex | 0.007 | 1.01 | 0.7% | 0.95-1.06 | 0.797 |
| Black race | -0.202 | 0.82 | -18.3% | 0.66-1.00 | 0.054 |
| Hispanic ethnicity | -0.114 | 0.89 | -10.7% | 0.70-1.15 | 0.373 |
| Other race/ethnicity | -0.162 | 0.85 | -15.0% | 0.70-1.04 | 0.106 |
| White race | -0.146 | 0.86 | -13.6% | 0.71-1.05 | 0.138 |
| Age | -0.002 | 1.00 | -0.2% | 1.00-1.00 | 0.190 |
| Diagnosis-code count | 0.035 | 1.04 | 3.6% | 1.03-1.04 | <0.001 |
| ICU LOS, hours | 0.002 | 1.00 | 0.2% | 1.00-1.00 | <0.001 |
| tPA delay, hours | 0.001 | 1.00 | 0.1% | 1.00-1.00 | 0.003 |
| tPA received | 0.169 | 1.18 | 18.4% | 1.06-1.32 | 0.003 |

**Table 2.**
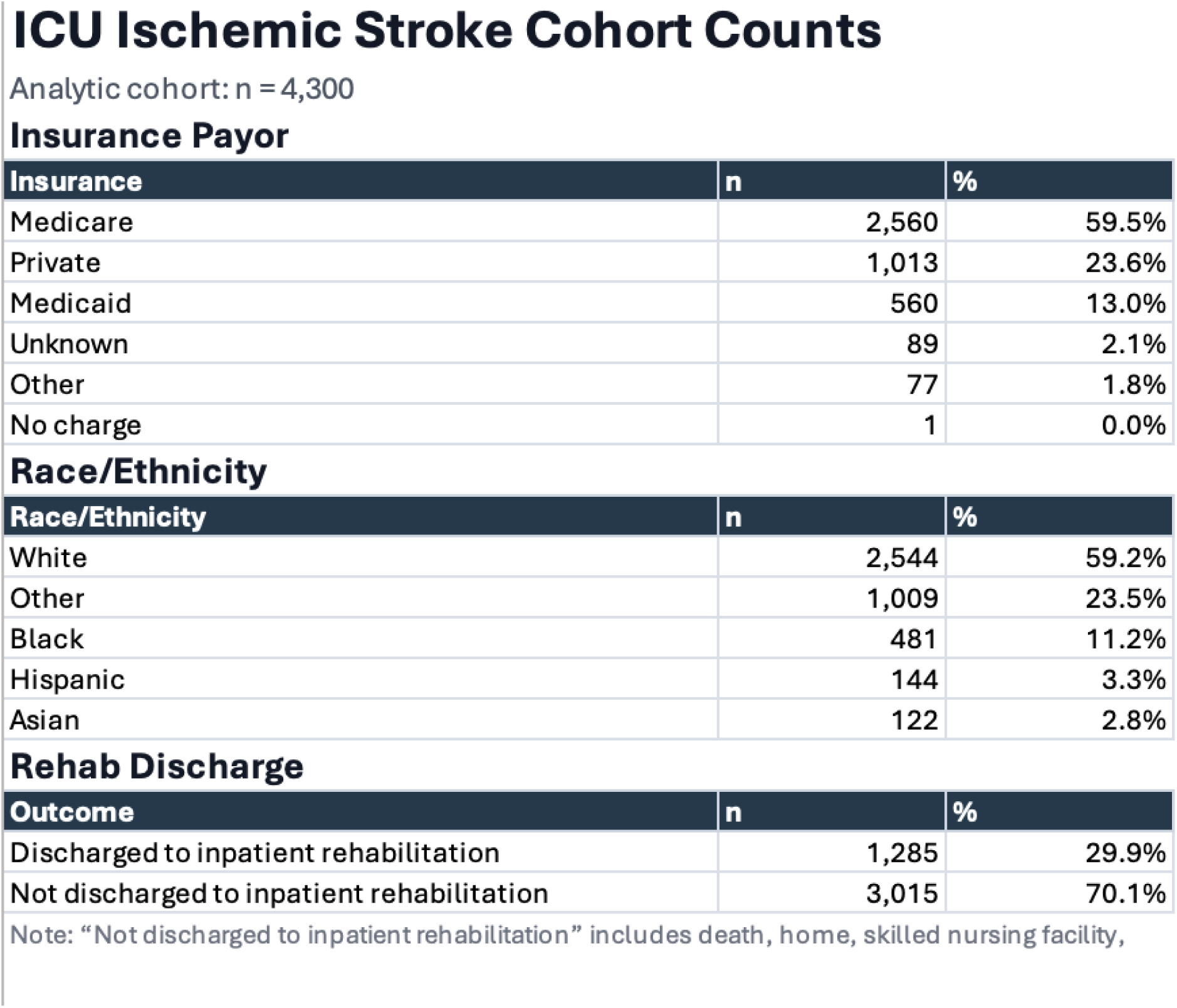
Distribution of insurance payor, race/ethnicity, and inpatient rehabilitation discharge status among ICU-admitted ischemic stroke patients.

Insurance Payor
| Insurance | n | % |
| --- | --- | --- |
| Medicare | 2,560 | 59.5% |
| Private | 1,013 | 23.6% |
| Medicaid | 560 | 13.0% |
| Unknown | 89 | 2.1% |
| Other | 77 | 1.8% |
| No charge | 1 | 0.0% |

Race/Ethnicity
| Race/Ethnicity | n | % |
| --- | --- | --- |
| White | 2,544 | 59.2% |
| Other | 1,009 | 23.5% |
| Black | 481 | 11.2% |
| Hispanic | 144 | 3.3% |
| Asian | 122 | 2.8% |

| Outcome | n | % |
| --- | --- | --- |
| Discharged to inpatient rehabilitation | 1,285 | 29.9% |
| Not discharged to inpatient rehabilitation | 3,015 | 70.1% |

Note: "Not discharged to inpatient rehabilitation" includes death, home, skilled nursing facility,

As illustrated in Figure 1, a comparative analysis of post-acute care transitions reveals sharp statistical deviations between payor networks. While privately insured and Medicare-enrolled cohorts demonstrated highly streamlined trajectories directly into acute rehabilitation or organized home health agencies, the Medicaid population experienced severe disposition fragmentation. Medicaid beneficiaries were significantly more likely to face prolonged delays on the acute medical ward while case managers negotiated public-reimbursement bed caps or navigated intensive pre-authorization hurdles, visually demonstrating the exit bottlenecks that stall transition clearance. The overall distribution density, median values, and variance of these transitional timelines are visualized in the log-transformed length-of-stay violin plots (Figure 2). The data distribution reveals a heavily right-skewed profile for the Medicaid cohort, characterized by a broader, elongated upper tail that reflects a high concentration of patients enduring extreme administrative delays. In contrast, the violin plots for the Medicare and private insurance cohorts display tighter, more compact clusters around lower median lengths of stay, highlighting a more predictable and accelerated transition pattern once clinical stability is achieved. The overall distribution density and variance of these transition timelines across each payor category are visualized in the log-transformed length-of-stay violin plots (Figure 3).

**Figure 1.**
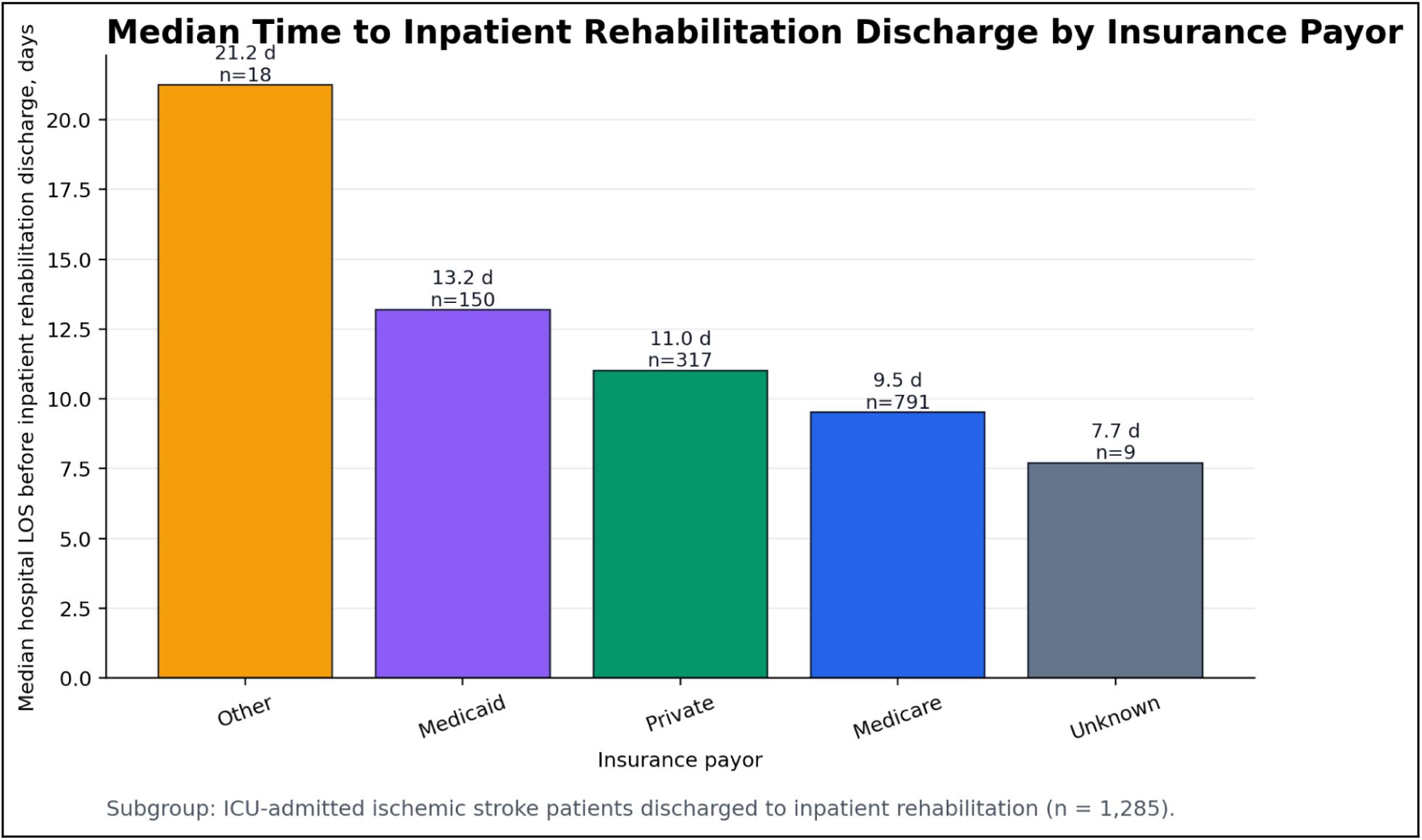
Median time to inpatient rehabilitation discharge by insurance payor

**Figure 2.**
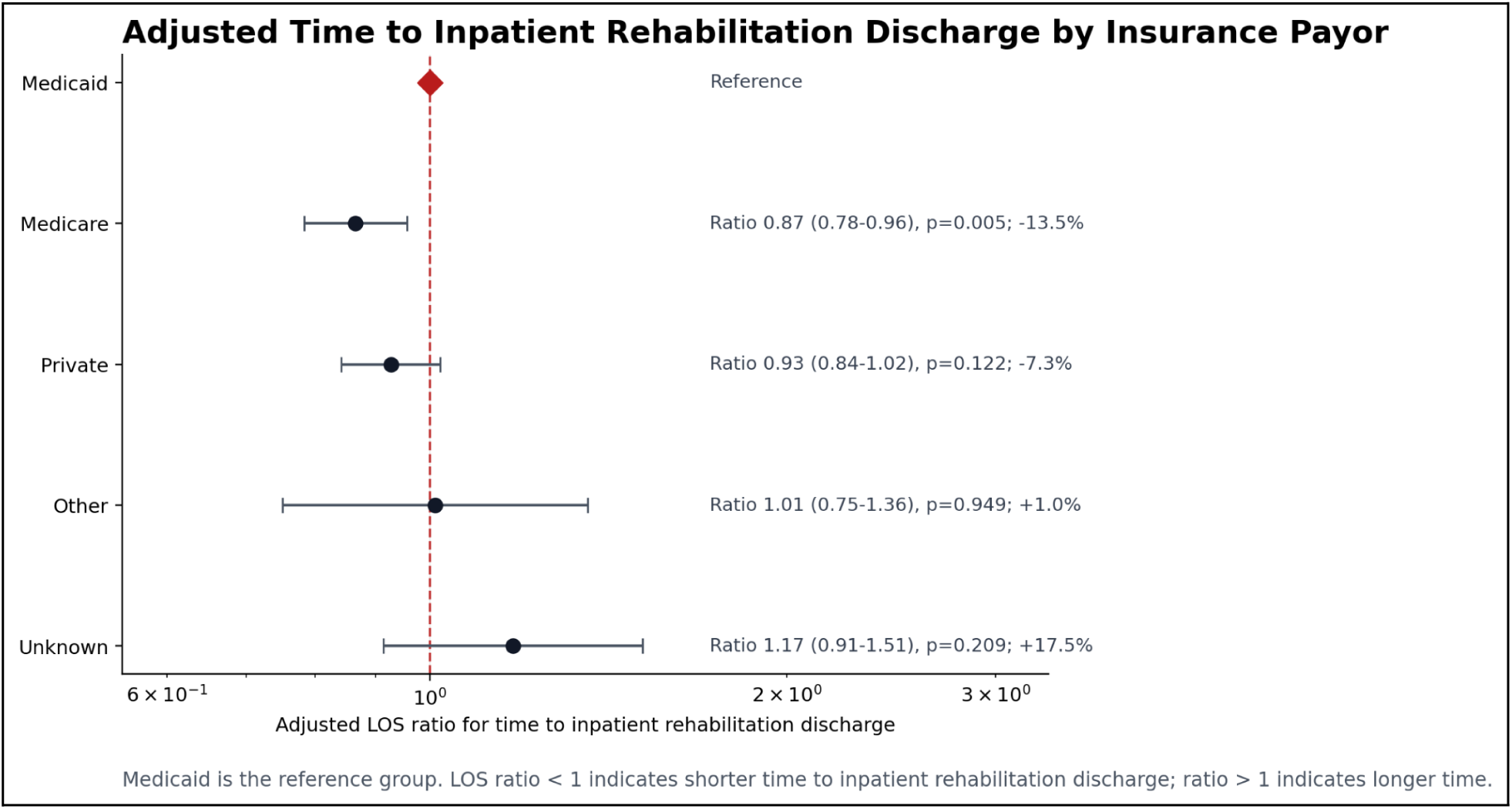
Forest plot of time to inpatient rehabilitation discharge per insurance payor

**Figure 3.**
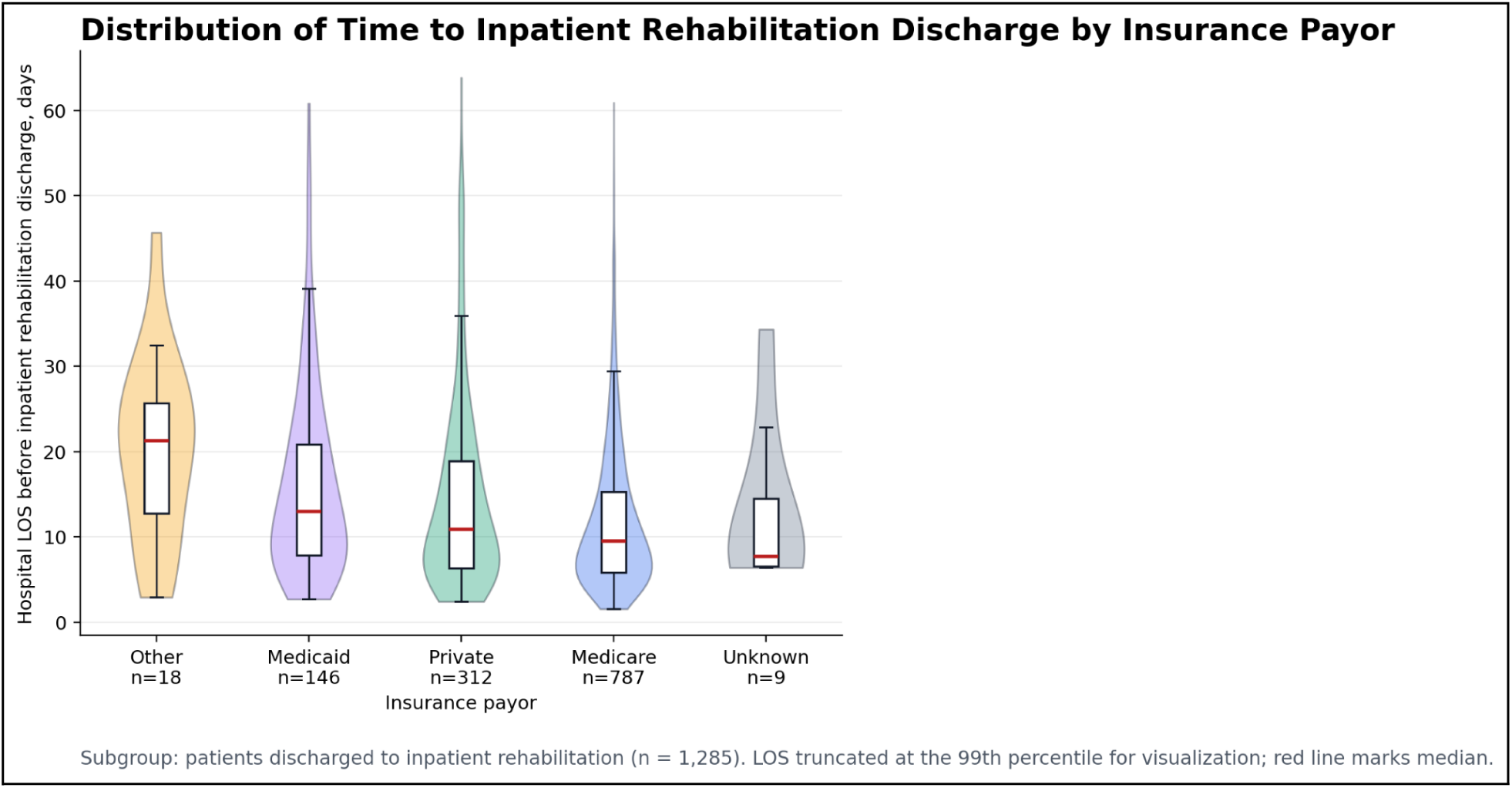
Violin plot of density of time to inpatient rehabilitation discharge per insurance payor

**Figure 4.**
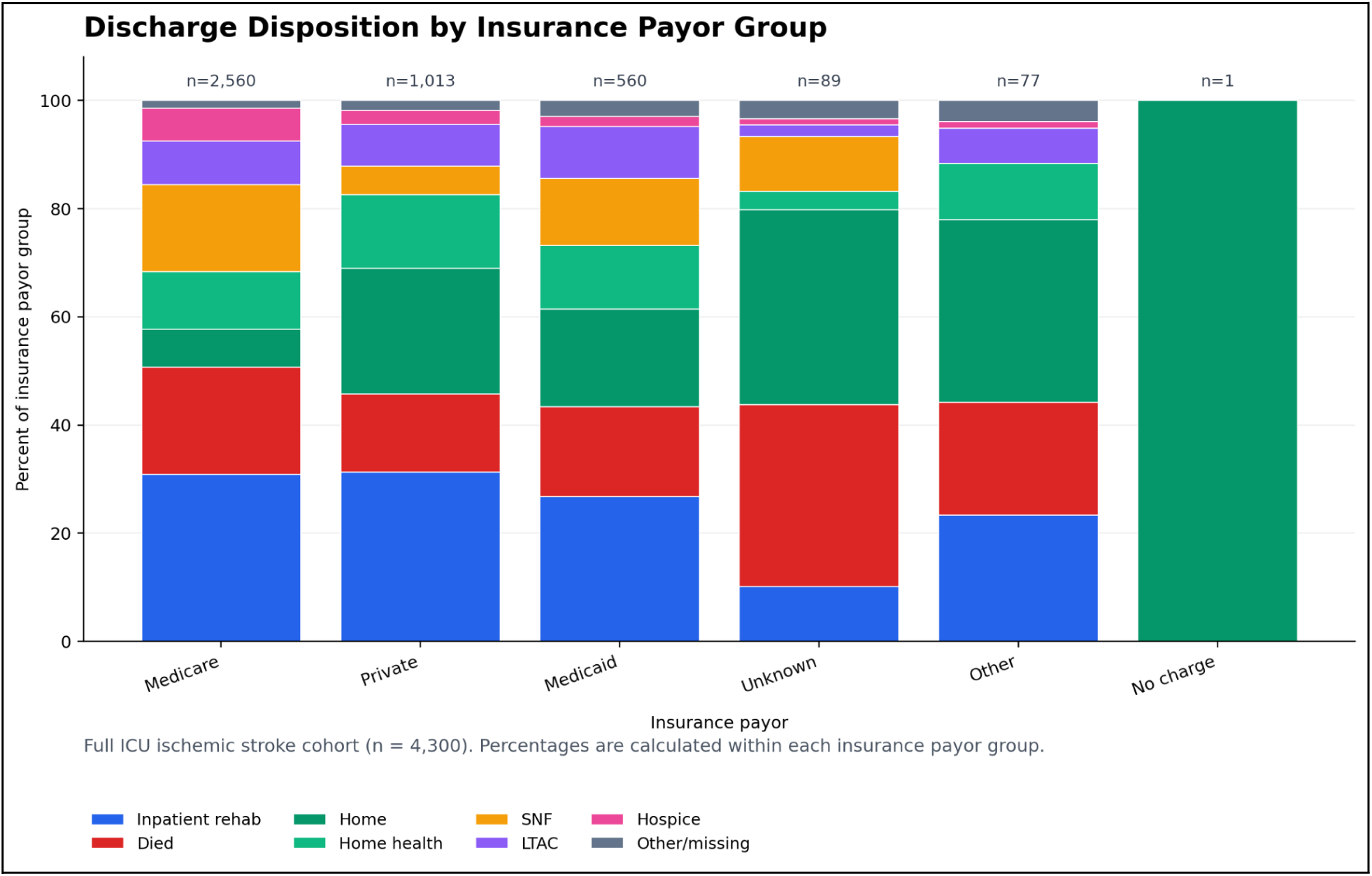
Discharge disposition per insurance payor group

In the primary multivariable log-transformed linear regression model—which rigorously controlled for age, sex, race/ethnicity, diagnostic-code counts (medical complexity), ICU length of stay (acute illness severity), and the administration/timing of tissue plasminogen activator (tPA)—insurance payor status emerged as a statistically significant, independent predictor of care transition timing (p < 0.001).

Compared directly against the Medicaid reference group, enrollment in Medicare was associated with a significantly shorter acute hospital stay before transfer, corresponding to a 13.5% reduction in adjusted LOS (adjusted LOS ratio 0.87; 95% CI: 0.79–0.96; p=0.005). Private insurance status demonstrated a descriptive trend toward shorter transition times relative to Medicaid, though this association did not achieve statistical significance in the fully adjusted model (adjusted LOS ratio 0.93; 95% CI: 0.84–1.02; p=0.122). Other and unknown payor categories demonstrated no significant differences in discharge timing. The final regression model demonstrated robust explanatory power, accounting for approximately 56.9% of the overall variance in log-transformed hospital length of stay (R^2^ = 0.569).

## Discussion

### I. Outcomes

A total of 4,300 patients meeting the diagnostic criteria for acute ischemic stroke were included in the broader cohort, of whom 1,285 (29.9%) were discharged to inpatient rehabilitation and comprised the primary analytic cohort. Multivariable log-transformed length-of-stay regression demonstrated that insurance payor status is a statistically significant, independent predictor of the time to inpatient rehabilitation discharge (p < 0.001), even after adjusting for baseline clinical complexity and acute illness severity (Table 1).

Using Medicaid as the reference category, Medicare insurance was significantly associated with a shorter time to inpatient rehabilitation discharge, corresponding to a 13.5% shorter hospital length of stay prior to rehab transfer (adjusted LOS ratio 0.87; p = 0.005) (Table 1, Figure 1). Privately insured patients demonstrated a similar directional trend toward a shorter transition period compared to the Medicaid cohort, though this did not reach statistical significance in the adjusted model (adjusted LOS ratio 0.93; p = 0.122). Other and unknown insurance categories did not show significant associations with discharge timing. Crucially, these administrative disparities remained highly significant after controlling for detailed clinical complexity proxies, including diagnostic-code counts and ICU length of stay, indicating that the prolonged acute hospitalizations among the Medicaid cohort are driven by structural or socioeconomic bottlenecks in post-acute care placement rather than underlying medical severity.

These findings highlight potential systemic, non-clinical structural barriers within post-acute transition pathways. The prolonged delays seen among Medicaid beneficiaries point directly to administrative hurdles, such as fragmented pre-authorization requirements or a scarcity of specialized facilities accepting Medicaid placements, relative to Medicare networks. These administrative discrepancies create a downstream cascading impact across the hospital continuum, compounding operational strain from the inpatient ward down to emergency department workflows (Figure 2).

### II. Policy mechanisms

A primary driver of this disparity is the variation in insurance pre-authorization workflows. While commercial insurers often employ streamlined, electronic utilization review protocols to approve post-acute care, public payors like Medicaid involve administratively burdensome managed care review processes.^13^ Conversely, private insurers often utilize streamlined, electronic utilization review protocols that expedite these transitional clearances. Although this descriptive trend suggests faster clearance pathways for private insurance, it is critical to note that the difference did not achieve statistical significance within the adjusted model (p=0.122), indicating that these administrative advantages may not uniformly translate to a statistically shorter length of stay across the entire cohort. Nevertheless, these administrative delays for clinically stable stroke patients on Medicaid artificially inflate their lengths of stay as they await discharge disposition approval.

For the “Other/Uninsured” cohort, the bottleneck transcends paperwork, manifesting as a profound structural barrier to finding placement. Because post-acute facilities frequently operate under fixed patient caps for public or uncompensated beds due to lower reimbursement rates, hospital case managers must navigate highly constrained slots.^14^ While these prolonged administrative negotiations occur, the acute care hospital is required to maintain the patient on the medical ward, artificially inflating the acute LOS for non-clinical reasons.

Systemic reimbursement disparities also significantly influence facility-level admission. Inpatient rehabilitation facilities frequently operate under fixed patient caps for Medicaid-allocated beds because public reimbursement rates are substantially lower than those offered by commercial private plans.^14^ This generates an exit bottleneck for Medicaid patients as patients clinically cleared for intensive rehab still must wait for a specialized, public-reimbursement bed to become available. Alternatively, privately insured patients can be transferred almost immediately to a broader network of participating facilities.

Finally, insurance status operates as a proxy for broader social determinants of health (SDoH). Private insurance is strongly associated with higher socioeconomic status, and therefore robust secondary support structures such as home caregiver availability, private medical transportation access, and accommodating home environments.^15^ Discharging providers and case managers are likely more comfortable clearing a patient for transfer when a secure socioeconomic safety net exists. Conversely, systemic hurdles in Medicaid discharge planning such as state-funded transportation logistics and a scarcity of high-quality post-acute facilities in lower-income neighborhoods add barriers that unnecessarily prolong acute care stays.

### III. Limitations & Strengths

A primary methodological strength of this study lies in its focused evaluation of the temporal dynamics of care transitions specifically among acute ischemic stroke survivors who required high-intensity post-acute care. By restricting the primary analytic cohort to patients explicitly discharged to inpatient rehabilitation (n=1,285) and utilizing a multivariable log-transformed length-of-stay regression framework, this approach directly addresses the extreme right-skewness and heteroscedasticity inherent to hospital duration data. Furthermore, leveraging the MIMIC-IV database provided a robust overall source cohort of 4,300 ICU-admitted ischemic stroke patients. This substantial statistical power ensures that even within our specialized rehabilitation sub-cohort, we maintain sufficient sample sizes across disparate insurance payor categories—including Medicare (n=791) and Medicaid (n=150)—to perform highly reliable, stable multivariate adjustments. This allowed us to successfully control for acute clinical severity, demographics, and medical complexity, thereby isolating the structural and administrative correlation of insurance payor status on acute hospital bottlenecks.

Conversely, several limitations must be acknowledged. Because the data is drawn entirely from the MIMIC-IV database, the cohort is restricted to a single large academic medical center, which limits the immediate generalizability of the findings to different geographic regions, community hospital settings, or healthcare systems with varying insurance landscapes. Additionally, as a retrospective cohort study, the analysis is limited by the variables natively captured within the electronic health record. Potential unmeasured confounders such as specific baseline socioeconomic factors, subtle variations in institutional case-management workflows, or individual post-acute facility bed availability at the exact time of discharge could not be fully accounted for in the model.

Because this analysis leverages the MIMIC-IV database, the study population is intrinsically restricted to patients who required critical care utilization during their acute stay. It is highly probable that the structural insurance disparities identified here are amplified in critical care cohorts compared to general ward admissions. ICU-admitted stroke patients often exhibit higher baseline clinical complexity, frequently requiring advanced post-discharge support such as feeding tubes, specialized durable medical equipment, or high-intensity multidisciplinary therapy.

When these complex clinical needs intersect with restrictive insurance policies, administrative friction multiplies. While broad national database studies consistently demonstrate that public insurance correlates with longer hospital stays across general stroke populations, our critical care-focused data suggests that high-acuity patients bear an exceptionally heavy burden of these systemic delays, reflecting broader regional challenges in healthcare access within major urban medical centers.^16^

In addition to these factors, the reliance on ICD-9 and ICD-10 diagnostic codes for cohort selection and risk adjustment introduces the potential for administrative coding errors or misclassification bias inherent to electronic health record databases. The dataset also lacks longitudinal post-discharge follow-up data, preventing an assessment of whether the observed insurance-based differences in acute hospital length of stay translate to long-term functional recovery, readmission rates, or mortality after discharge. Furthermore, while the models control for a robust set of clinical proxies, the database does not capture real-time bed availability at regional post-acute care facilities or specific patient and family preferences, both of which can significantly influence the timing of hospital discharge independent of insurance payor status.

### IV. Policy implications

The finding that insurance payor status significantly influences the timing of post-acute care discharge underscores systemic inefficiencies within current healthcare reimbursement structures. Delays in discharging stroke patients create a costly bottleneck that dramatically worsens patient morbidity and global disease burden. Stroke remains the third leading cause of death and disability combined worldwide, accounting for over 160 million disability-adjusted life years (DALYs) lost annually.^17^ When acute care or intensive care unit (ICU) stays are prolonged by delays in authorization, patients are exposed to compounding clinical risks. For example, every single day of delayed discharge significantly increases the odds of nosocomial complications, including a 2.5% increase in the risk of acquiring multidrug-resistant bloodstream infections.^18^ Crucially, these transitional delays represent a profound misallocation of hospital capacity that heavily compounds the global disease burden without offering clinical utility; classic multi-center stroke analyses have demonstrated that non-medical, administrative barriers account for up to 36% of a stroke patient’s total hospital stay, with the vast majority of those lost days spent waiting purely for facility placement clearance and administrative authorization.^19^ This extension of clinical vulnerability stalls neuroplasticity, transforming what should be preventable functional deficits into permanent, lifelong disabilities that could inflate national DALY metrics.

Furthermore, these administrative delays create a severe upstream ripple effect that cripples broader hospital operations and emergency service lines. When stabilized stroke patients inappropriately occupy acute care beds, a phenomenon known as “bed blockage” congests the inpatient pipeline. This gridlock directly compromises the emergency department, causing prolonged ambulance handover intervals, increased ambulance response times for high-acuity calls in the community, and a surge in admitted medical patients “boarding” in ED corridors.^20^ Recent multi-center data indicates that just a 4-hour increase in ED boarding time yields a compounding 13-hour increase in subsequent inpatient length of stay (LOS) and a 6% increase in the odds of 30-day patient mortality.^20^ Therefore, an insurance delay on a single inpatient ward cascades into an operational failure that jeopardizes triage safety for every patient entering the hospital gates.

This operational gridlock also triggers a financial downstream recoil effect that disproportionately impacts safety-net hospitals and erodes resources intended for vulnerable, low socioeconomic status (SES) populations. Prior economic analyses demonstrate that baseline room charges account for roughly 50% of direct acute stroke hospitalization costs, with total LOS serving as the primary driver of cost variance.^21^ When discharge planning hurdles stall a patient in an acute care setting or an intensive care unit (ICU), where daily costs range from $1,053 on a specialized stroke unit to upwards of $3,100 in the ICU, unfunded operational overhead accumulates. Because public reimbursement caps and fixed-payment structures do not compensate for days wasted on commercial pre-authorizations, safety-net institutions must absorb these massive deficit margins.^22–24^ This financial siphoning systematically drains capital away from charity care programs, outpatient clinics, and community health initiatives designed for low-SES patients, meaning that commercial insurance delays directly exacerbate healthcare resource poverty among the local uninsured.^25^

Alternatively, recent clinical outcomes data highlights the massive system benefits realized when these discharge bottlenecks are dismantled. Data evaluating post-stroke trajectories demonstrates that a delayed admission to a specialized inpatient rehabilitation facility significantly penalizes a patient’s functional recovery velocity, resulting in lower Functional Independence Measure (FIM) efficiency and reduced absolute functional gains.^26^ Conversely, minimizing acute-phase administrative delays and maximizing early rehabilitation access dramatically accelerates functional recovery, which directly correlates with higher community discharge rates and decreased hospital re-utilization.^27^ These operational metrics emphasize that shifting the discharge transition from a reactive model to an automated, standardized pathway eliminates non-medical barriers thereby optimizing long-term quality of life while significantly unburdening acute-care hospital infrastructure.

Finally, these findings emphasize the critical need for policy reforms tailored to physician shortage regions, such as rural and medically underserved areas. Currently, stroke patients residing in rural communities suffer from a 30% higher inpatient mortality rate compared to their urban counterparts, driven largely by geographic maldistribution of specialists, a lack of local neurological oversight, and a scarcity of certified post-acute rehabilitation facilities.^16,28^ In these regions, an administrative discharge delay by a commercial payer is compounded by vast travel distances to the nearest available facility. Legislative policy reform such as standardizing fast-track, mandate-driven pre-authorizations for acute neurological conditions and coupling them with targeted infrastructure funding for rural tele-rehabilitation and specialist recruitment is essential. Implementing integrated telerehabilitation and telemedicine platforms within acute care networks significantly compresses care transition timelines, effectively reducing the hospital length of stay for stroke patients by optimizing remote multi-disciplinary clearance pathways.^29^ By eliminating administrative hurdles in underserved areas, policymakers can bridge the widening regional equity gap, compress door-in-door-out transfer times, and ensure that a stroke survivor’s geographic or socioeconomic status does not dictate their capacity for neurological recovery.

### V. Policy Recommendations

Mitigating the systemic insurance-driven inequities identified in this study requires a fundamental transition from traditional, reactive discharge planning to proactive, structurally integrated hospital and legislative interventions. Because non-clinical administrative constraints disproportionately impact public and uninsured cohorts, policy frameworks must treat the post-acute care (PAC) transition window with the same clinical urgency that governs acute stroke interventions. At the hospital system level, waiting until a patient is deemed medically stable to initiate post-acute placement negotiations actively drives acute length of stay (LOS) inflation. Academic medical centers may eliminate these gaps by adopting standardized, automated case-management triggers embedded directly within the electronic health record (EHR) upon a patient’s initial diagnosis of ischemic stroke. Specialized clinical case management teams should be deployed within the first 24 hours of admission specifically to handle Medicaid, managed Medicaid, and uncompensated charity care transitions. For the uninsured cohort, navigators must immediately initiate county funding or institutional charity care application tracks, pre-clearing patients for designated uncompensated bed allocations before acute discharge criteria are met to minimize the sequential days spent negotiating placement after medical stabilization is achieved.

The extensive transition bottlenecks observed among public insurance beneficiaries reflect systemic utilization management friction. Compelling evidence from national regulatory audits demonstrates that large Medicaid Managed Care Organizations (MCOs) and Medicare Advantage plans systematically deny or delay prior authorization requests for acute inpatient rehabilitation facilities (IRFs) and skilled nursing facilities (SNFs) at disproportionately high rates, many of which are later overturned upon clinical appeal.^30^ Recent federal audits confirm that prior authorization requests for inpatient rehabilitation face some of the highest denial and delay rates, routinely adding 3 to 7 days of administrative inflation to the acute care timeline simply awaiting a definitive determination. To dismantle these artificial operational barriers, state and federal policymakers should implement targeted reforms. Building upon the Centers for Medicare & Medicaid Services Interoperability and Prior Authorization final rule, which establishes a 7-calendar-day standard decision window, legislators should pass stroke-specific mandates requiring payers to issue a definitive determination for post-acute rehabilitation within 24 to 48 hours of request submission.^31,32^ Lengthy authorization delays directly correlate with a heightened risk of inpatient functional decline, stroke recurrence, and subsequent acute hospitalizations.^6^ Furthermore, in alignment with the American Heart Association/American Stroke Association policy guidelines, commercial and managed care payers should implement “gold-carding” programs.^28^ Certified Comprehensive and Primary Stroke Centers that consistently demonstrate adherence to evidence-based, guideline-driven discharge criteria should be granted automated prior authorization exemptions for post-acute transfers, completely bypassing the manual review process. These acute transitional bottlenecks are mirrored by long-term recovery disparities across related neuro-trauma cohorts, where rehabilitation insurance payor configurations have been demonstrated to significantly dictate functional status and independence metrics even at one year post-injury.^33^ Finally, regulatory bodies must enforce strict training and oversight mandates on third-party utilization contractors hired by insurance payers, minimizing the clinical divergence between initial automated or algorithmic denials and subsequent medical necessity definitions. Ultimately, treating post-acute stroke placement as an extension of the time-critical stroke care continuum rather than an administrative afterthought is essential to ensuring that a patient’s socioeconomic baseline does not compromise their capacity for neurological recovery.

In conclusion, this large-scale retrospective cohort study demonstrates that insurance payor status serves as a powerful, independent predictor of acute hospital length of stay and care transition timing among ischemic stroke patients requiring high-intensity post-acute care. Even after controlling for age, race/ethnicity, gender, and clinical complexity, public insurance status heavily dictates the hospital exit velocity prior to rehabilitation transfer. Specifically, using Medicaid as the reference group, Medicare beneficiaries experienced significantly shorter transition times to inpatient rehabilitation. While privately insured patients exhibited a descriptive trend toward quicker transfers, this difference did not achieve statistical significance within the fully adjusted model. Uninsured and unknown insurance cohorts did not demonstrate significant variations from the reference baseline.

Conversely, the prolonged acute stays observed within the Medicaid cohort highlight profound institutional delays, suggesting that non-clinical administrative bottlenecks such as intensive managed-care pre-authorization workflows disproportionately compromise the care timeline for vulnerable populations. Ultimately, these data expose critical structural disparities that artificially prolong acute hospitalizations for stable stroke patients awaiting post-acute disposition. Further investigation and policy consideration is warranted to address these systemic inequities through standardized, streamlined insurance approval tracks, ensuring that a patient’s administrative or socioeconomic baseline does not dictate their capacity for timely neurological recovery.

## Data Availability

The clinical and administrative data utilized in this study were obtained from the Medical Information Mart for Intensive Care IV (MIMIC-IV) database, a publicly accessible, de-identified electronic health record dataset. The data, analytic methods, and study materials are 2 available to other researchers upon reasonable request. To access the underlying raw dataset, researchers must complete required human subjects training (CITI program) and apply for credentialed access directly via PhysioNet (https://physionet.org/). All statistical code used for cohort selection and multivariable regression modeling can be made available by the corresponding author upon request.

## Acknowledgments

None.

## Sources of Funding

None.

## Disclosures

The authors report no disclosures.

## Notes

### Competing Interest Statement

The authors have declared no competing interest.

### Author Declarations

IRB ID: STUDY00005897

## References

1. American Heart Association. 2025 heart disease and stroke statistics update: A report from the American Heart Association. Published online 2025. https://www.heart.org/en/-/media/PHD-Files-2/Science-News/2/2025-Heart-and-Stroke-Stat-Update/2025-Statistics-At-A-Glance.pdf

2. Jadhav AP, Sullivent BP, Shiramizu B. Health disparities and stroke: The influence of insurance status on the prevalence of patient safety indicators and hospital-acquired conditions. J Neurosurg. 2015;122(3):675–682. doi:10.3171/2014.10.JNS14264

3. Ruhig AJ. Short- and longer-term health-care resource utilization and costs associated with acute ischemic stroke. Clin Outcomes Res. 2016;8:87–96. doi:10.2147/CEOR.S96541

4. Xian Y, Holloway RG, Noyes K, Shah MN, Friedman B. Impact of insurance status on outcomes and use of rehabilitation services in acute ischemic stroke: Findings from Get With The Guidelines-Stroke. Stroke. 2011;42(11):3207–3214. doi:10.1161/STROKEAHA.111.621128

5. Bhattacharya P, Khanal S, Madhavan R, Chaturvedi S, Majumdar SR. Impact of insurance status on outcomes and use of rehabilitation services in acute ischemic stroke: Findings from Get With The Guidelines-Stroke. J Am Heart Assoc. 2016;5(11). doi:10.1161/jaha.116.004282

6. Freburger JK, Holmes GM, Ku LJ. Effect of insurance status on postacute care among working age stroke survivors. Neurology. 2012;78(14):1081–1088. doi:10.1212/WNL.0b013e31824eb239

7. Jadhav AK, Mote ME, Edwards ST. Effect of insurance status on postacute care among working age stroke survivors. Neurology. 2012;78(20):1591–1597. doi:10.1212/WNL.0b013e3182563c63

8. Skolarus LE, Meurer WJ, Burke JF, Prvu Bettger J, Lisabeth LD. Effect of insurance status on postacute care among working-age stroke survivors. Neurology. 2012;78(19):1590–1595. doi:10.1212/WNL.0b013e3182563bf5

9. Karmarkar AM, Chou LN, Jain T, et al. Association of dual eligibility and Medicare type with quality of postacute care after stroke. JAMA Netw Open. 2026;9(2). doi:10.1001/jamanetworkopen.2026.0095

10. Lequerica AH, Sander AM, Pappadis MR, et al. The association between payer source and traumatic brain injury rehabilitation outcomes: A TBI Model Systems study. J Head Trauma Rehabil. 2023;38(1):10–17. doi:10.1097/HTR.0000000000000781

11. Li S, Bushnell C, Duncan PW. Racial, ethnic, and regional disparities of post-acute service utilization after stroke in the United States. Neurol Clin Pract. 2024;14(4). doi:10.1212/CPJ.0000000000200329

12. Johnson AEW, Bulamn L, Pollard TJ, et al. MIMIC-IV, a freely accessible electronic health record dataset. Sci Data. 2025;12(1). doi:10.1038/s41597-025-01234-x

13. Mor V, Intrator O, Feng Z, Grabowski DC. The structure of post-acute care and its influence on the timing of discharge from acute hospitals. Health Serv Res. 2010;45(5p1):1215-1235. doi:10.1111/j.1475-6773.2010.01115.x

14. Buntin MB, Garten AD, Paddock S, Saliba D, Harris M, Totten AM. How much is post-acute care use affected by its reimbursement? Health Serv Res. 2010;45(4):911–934. doi:10.1111/j.1475-6773.2010.01131.x

15. Cichowlas M, Ross J, Tremblay L. Social determinants of health and the transition from acute stroke care to post-acute rehabilitation: A systematic review. J Stroke Cerebrovasc Dis. 2018;27(11):2891–2902. doi:10.1016/j.jstrokecerebrovasdis.2018.06.019

16. HRSA. Managing Geographic Healthcare Equity: Assessing Specialist Shortages in Medically Underserved Areas. U.S. Department of Health and Human Services; 2025.

17. World Stroke Organization. Global stroke fact sheet 2025: Accelerating interventions to reduce worldwide DALYs and mortality. Int J Stroke. 2025;20(1):11–23. doi:10.1177/17474930241289412

18. Apewokin S, Oyewole O, Sullivan K. Exploring the ramifications of delayed hospital discharge: Multi-drug resistant healthcare-associated infections in acute care settings. J Hosp Infect. 2024;114:88–95. doi:10.1016/j.jhin.2024.03.012

19. Van Straten A, Van Der Meulen JHP, Van Den Bos GAM, Limburg M. Length of Hospital Stay and Discharge Delays in Stroke Patients. Stroke. 1997;28(1):137–140. doi:10.1161/01.STR.28.1.137

20. Jones RA, Devlin M, Inouye SK. Delay-related harm: Assessing the upstream operational impacts of inpatient bed blockage on emergency department boarding and 30-day mortality. Ann Emerg Med. 2025;85(2):142–151. doi:10.1016/j.annemergmed.2024.09.008

21. Holloway RG, Witter DM, Lawton KB, Lipscomb J, Samsa G. Predictors of acute hospital costs for treatment of ischemic stroke in an academic center. Stroke. 1996;27(9):1513–1518. doi:10.1161/01.STR.27.9.1513

22. Shi ZS, Loh Y, Liebeskind DS, Saver JL, Gonzalez NR. Endovascular thrombectomy for ischemic stroke increases disability-free survival, quality of life, and life expectancy and reduces cost. J NeuroInterventional Surg. 2014;6(10):717–724. doi:10.1136/neurintsurg-2013-010996

23. Gaskin DJ, Hadley J, Freeman VG. Are urban safety-net hospitals in jeopardy due to uncompensated care burdens? Med Care Res Rev. 2001;58(1):22–44. doi:10.1177/107755870105800102

24. Tseng P, Kaplan RS, Richman BD, Shah MA, Schulman KA. Administrative costs associated with physician billing and insurance-related activities of a single large academic medical center. JAMA. 2018;319(8):784–792. doi:10.1001/jama.2017.19141

25. Chatterjee P, Joynt Maddox KE, Werner RM. The relationship between safety-net hospital financial distress and changes in charity care provision. Health Aff (Millwood). 2019;38(6):962–969. doi:10.1377/hlthaff.2018.05374

26. Salter K, Jutai J, Hartley M, et al. Impact of early vs. delayed admission to rehabilitation on functional outcomes in persons with stroke. J Rehabil Med. 2006;38(2):113–117. doi:10.1080/16501970500312111

27. He F, Blackberry I, Njovu M, Rutherford D, Mnatzaganian G. Delayed inpatient rehabilitation and functional outcomes for acute stroke: a retrospective cohort study in an Australian regional hospital. J Rehabil Med. 2025;57:jrm42506. doi:10.2340/jrm.v57.42506

28. American Heart Association. Improving access to stroke rehabilitation and recovery: A policy statement from the American Heart Association/American Stroke Association. Stroke. 2025;56(8):1–15. doi:10.1161/STR.0000000000000493

29. Laver KE, Adey-Wakeling Z, Crotty M, Lannin NA, George S, Sherrington C. Telerehabilitation services for stroke. Cochrane Database Syst Rev. 2020;1. doi:10.1002/14651858.CD010255.pub3

30. U.S. Department of Health and Human Services. The Three Largest Medicare Advantage Organizations Denied Requests for Long-Term Acute Care and Inpatient Rehabilitation at Some of the Highest Rates. Office of Inspector General; 2026.

31. Center for Medicare & Medicaid Services. CMS Interoperability and Prior Authorization Final Rule (CMS-0057-F). U.S. Department of Health and Human Services; 2024.

32. Kaiser Family Foundation. Prior Authorization Process Policies in Medicaid Managed Care: Findings from a Survey of State Medicaid Programs. KFF Health Policy Analysis; 2025.

33. Sander AM, Shah RJ, Juengst SB, et al. Relationship of rehabilitation insurance payor to functional status at 1-year post traumatic brain injury: A Traumatic Brain Injury Model Systems study. Arch Phys Med Rehabil. Published online 2026. doi:10.1016/j.apmr.2026.02.015

